# Cross-sectional association of skeletal muscle mass and strength with dietary habits and physical activity among first-year university students in Japan: Results from KEIJI-U study

**DOI:** 10.1101/2022.04.28.22274428

**Authors:** Toru Kusakabe, Hiroshi Arai, Yuji Yamamoto, Kazuwa Nakao, Yasunori Akamatsu, Yuki Ishihara, Tetsuya Tagami, Akihiro Yasoda, Noriko Satoh-Asahara

**Affiliations:** Department of Endocrinology, Metabolism, and Hypertension Research, Clinical Research Institute, National Hospital Organization Kyoto Medical Center, Kyoto, Japan; Health Administration Center, Kyoto Institute of Technology, Kyoto, Japan; Health and Medical Services Center, Shiga University, Shiga, Japan; Medical Innovation Center, Kyoto University Graduate School of Medicine, Kyoto, Japan; Department of Endocrinology and Metabolism, National Hospital Organization Kyoto Medical Center, Kyoto, Japan; Clinical Research Institute, National Hospital Organization Kyoto Medical Center, Kyoto, Japan

**Keywords:** sarcopenia, skeletal muscle mass, muscle strength, dietary habit, physical activity

## Abstract

**Background:** Recently, the high prevalence of young Japanese people who are underweight has received attention because of its potential impact on healthy life expectancy. Sarcopenia, defined as the presence of low muscle mass and function, is often observed in the underweight. However, few reports have described the prevalence and characteristics of sarcopenia in youth.

**Methods:** In this cross-sectional study, we measured skeletal muscle mass using a multifrequency bioelectrical impedance analysis device and handgrip strength (HGS) and administered questionnaires on dietary habits and physical activity to 1,264 first-year university students aged 18-20 years (838 males and 426 females). Sarcopenia was diagnosed based on the presence of both low skeletal muscle mass (SMI) and weak HGS according to the Asian Working Group for Sarcopenia.

**Results:** A total of 145 males (17.3%) and 69 females (16.2%) were diagnosed as underweight. Low SMI was diagnosed in 75 males (8.9%) and 18 females (4.2%), and weak HGS was diagnosed in 28 males (3.3%) and 10 females (2.3%). Then, sarcopenia was diagnosed in 8 males (1.0%) and 5 females (1.2%). There was a significantly higher prevalence of low SMI and/or weak HGS in underweight individuals compared with individuals in the other BMI ranges. The multivariate analyses indicated that SMI and HGS were significantly associated with BMI in both sexes. Furthermore, after adjusting for BMI, both SMI and HGS were significantly associated with physical activity in males, and SMI was significantly associated with energy intake and frequency of breakfast intake in females.

**Conclusions:** First-year university students showed a high incidence of being underweight with low SMI and/or weak HGS, but the prevalence of sarcopenia was low in both sexes. BMI, energy intake, frequency of breakfast intake, and physical activity were independently associated with skeletal muscle mass and strength in youth.

## Introduction

According to the 2019 National Health and Nutrition Survey in Japan, 3.9% of males and 11.5% of females are underweight (BMI < 18.5 kg/m^2^) [1]. Notably, this percentage is almost double (20.7%) among females in their twenties. There is concern not only about the loss of body fat but also regarding the loss of muscle mass in underweight individuals. Loss of muscle mass at a youthful age can lead to the development of sarcopenia and frailty in the future. Furthermore, the loss of muscle mass is considered a risk factor for type 2 diabetes later in life because skeletal muscle is the largest glucose-processing organ in the body [2, 3]. Being underweight in young females is also associated with a risk of osteoporosis, fractures, and giving birth to low-birth-weight infants with a high risk of developing cardiovascular and metabolic diseases [4-6].

Sarcopenia was proposed in 1989 to describe the age-related decrease of muscle mass [7]. According to the revised European Working Group on Sarcopenia in Older People (EWGSOP) guideline, probable sarcopenia is identified by low muscle strength, and diagnosis of sarcopenia is confirmed by additional documentation of low muscle quantity or quality. Furthermore, when low muscle strength, low muscle quantity/quality and low physical performance are all detected, sarcopenia is considered severe [8]. In 2014, the Asian Working Group for Sarcopenia (AWGS) published a consensus report describing the original cutoff values for Asian populations [9] and updated it in 2019 [10]. Muscle loss when no other cause is evident but aging itself is called “primary sarcopenia”, and muscle loss when one or more other causes are evident (i.e., activity-related, disease-related, or nutrition-related) is called “secondary sarcopenia” [8, 11]. Therefore, secondary sarcopenia can be observed even in young people.

Skeletal muscle mass and strength increase with growth throughout youth and young adulthood, reaching maximal levels up to ∼40 years of age before declining [12, 13]. Young people are more likely to increase their muscle mass under the influence of diet and exercise than would the elderly [14, 15]. Therefore, it is desirable to increase muscle mass as much as possible from a youthful age to avoid developing sarcopenia in the future. However, extreme calorie restriction due to a desire to lose weight is often observed among young people. Furthermore, according to the 2019 National Health and Nutrition Survey in Japan, only 28.4% of men and 12.9% of women in their twenties have a habit of exercising; these proportions are both lower than the overall rate [1]. These facts are undesirable for young people to increase muscle mass.

Thus far, there have been few studies on sarcopenia in large groups of young subjects. In this study, we conducted a survey on skeletal muscle mass and strength, dietary habits, and physical activity to investigate the prevalence of sarcopenia and examine factors associated with skeletal muscle mass and strength among first-year university students in Japan.

## Methods

### Study participants

A total of 1,287 first-year university students (850 males and 437 females) aged 18-20 years who underwent the health examination at either the Kyoto Institute of Technology or Shiga University in April 2019 were enrolled in this study (KEIJI-U study). Twenty-three subjects (12 males and 11 females) were excluded because of incomplete data, and finally 1,264 subjects (838 males and 426 females) were analyzed (Fig 1). Consent to participate was obtained from all participants before study inclusion. This study was reviewed and approved by the Ethics Committee of National Hospital Organization Kyoto Medical Center (approval number: 18-106) and was conducted in compliance with the ethical principles stated in the Declaration of Helsinki.

**Fig 1.**
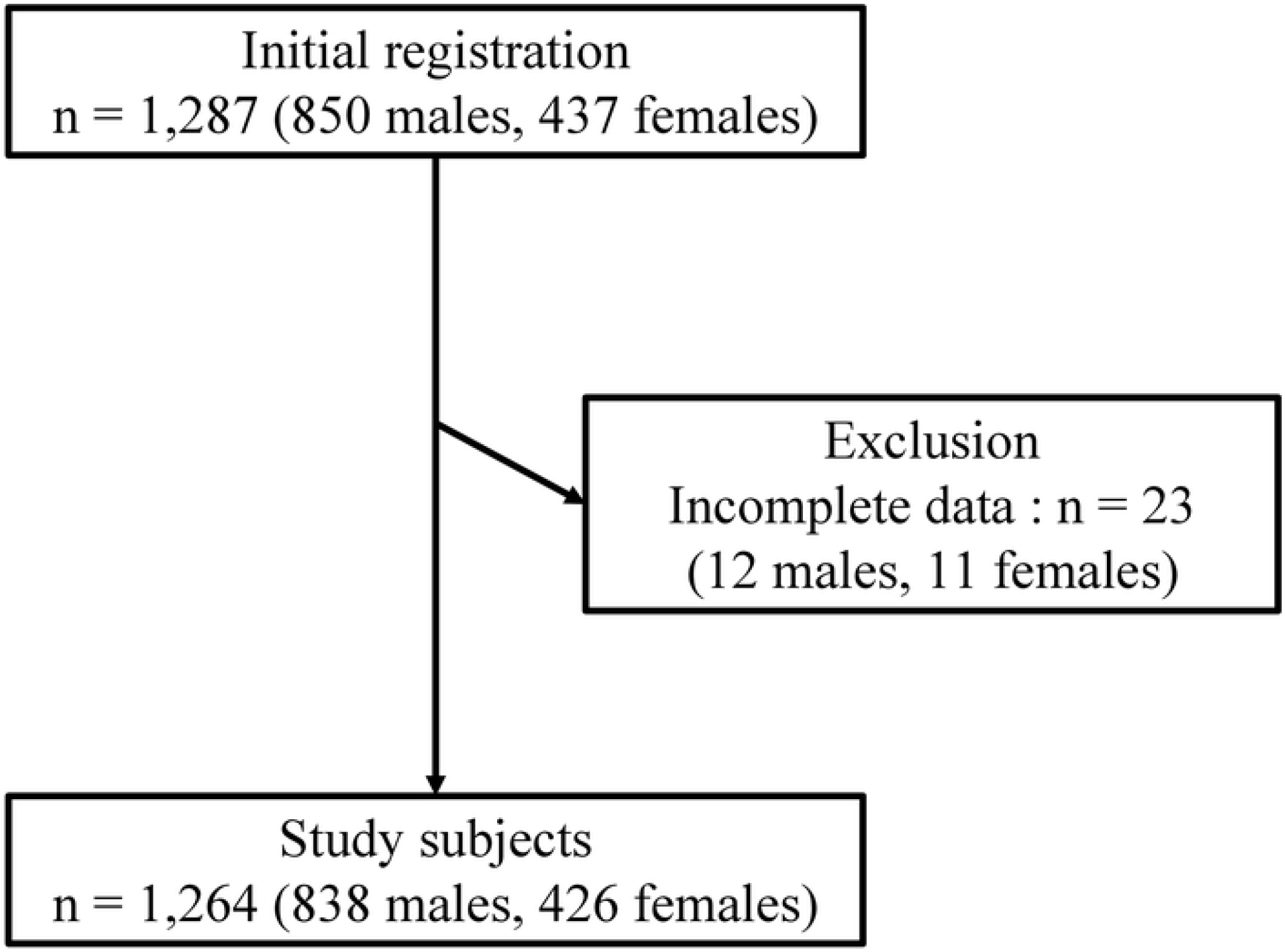
Flow chart of study subjects.

### Measurement of body composition and muscle strength

Body weight, total fat mass, body fat percentage, total muscle mass, appendicular skeletal muscle mass (ASM), total bone mass, and body water content were measured using a multifrequency bioelectrical impedance analysis (BIA) device (MC-780A-N, TANITA, Tokyo, Japan). An estimation formula for ASM in this model has been published, and a previous validation study demonstrated that body composition measured using this device was highly correlated with that measured using dual-energy X-ray absorptiometry measurements [16].

Muscle strength was measured as handgrip strength (HGS) using the Smedley grip force system (Grip-D, Takei Scientific Instruments Co, Ltd., Niigata, Japan) while the subject was in a standing position. Maximum isometric grip strength was assessed from two attempts for each hand.

### Assessment of dietary habits

The brief-type self-administered diet history questionnaire (BDHQ) was used to assess dietary habits [17, 18]. Briefly, the intake of 58 food and beverage items was evaluated to estimate energy intake (kcal/day), total/animal/vegetable protein intake (g/day), total/animal/vegetable fat intake (g/day), and carbohydrate intake (g/day) using a computer algorithm based on the Standard Tables of Food Composition in Japan [19]. Frequency of breakfast intake was scored as follows (1–9 points); 1: no breakfast, 2: less than 1 time/week, 3: 1 time/week, 4: 2 times/week, 5: 3 times/week, 6: 4 times/week, 7: 5 times/week, 8: 6 times/week, and 9: every morning.

### Assessment of physical activity

The International Physical Activity Questionnaire (IPAQ) short version was used to assess physical activity [20, 21]. Briefly, physical activity was calculated based on the time spent performing walking, moderate-, and vigorous-intensity activity. The metabolic equivalent (MET) levels were 3.3 for walking, 4.0 for moderate-intensity activity, and 8.0 for vigorous-intensity activity, respectively. Each physical activity was assessed as follows: each MET level × minutes of activity/day × days/week. Then, total physical activity (MET-minutes/week) was calculated as the sum of the walking, moderate-, and vigorous-intensity activities.

### Definitions of body physique

Body mass index (BMI, kg/m^2^) was calculated as body weight (kg) divided by height (m) squared. A BMI less than 18.5 kg/m^2^ was defined as underweight, a BMI of 18.5-24.9 kg/m^2^ was defined as normal, and a BMI of 25 kg/m^2^ or greater was defined as obesity based on the definition of obesity recommended by the Japan Society for the Study of Obesity.

### Definitions of sarcopenia

The skeletal muscle mass index (SMI, kg/m^2^) was calculated as the ASM divided by height (m) squared. Low skeletal muscle mass was defined as an SMI less than 7.0 kg/m^2^ for males and less than 5.7 kg/m^2^ for females when measured by BIA, and weak grip strength was defined as HGS less than 28 kg for males and less than 18 kg for females based on the criteria of the AWGS 2019 [10]. Sarcopenia was diagnosed based on the presence of both low skeletal muscle mass and weak grip strength.

### Statistical analyses

In this cross-sectional study, the data are expressed as the mean ± standard deviation. The correlations between the SMI or HGS and other factors were first analyzed by a simple linear regression analysis and then by a multiple linear regression analysis. The prevalence of low SMI and/or weak HGS was compared among the three groups (underweight, normal, and obesity) using the χ^2^-test. Then, as a post hoc test, multiple comparisons were performed using Tukey’s WSD test. The statistical analyses were performed using BellCurve for Excel version 3.20 (Social Survey Research Information Co., Ltd., Tokyo, Japan), and a two-sided *P* < 0.05 was considered statistically significant.

## Results

### Body physique of Japanese university students

The characteristics of the study subjects are summarized in Table 1. The average BMIs in males and females were 21.2 ± 3.0 and 20.5 ± 2.3 kg/m^2^, respectively, and both values were within the normal ranges. The BMI distributions in males and females are shown in Fig 2A. A total of 145 males (17.3%) and 69 females (16.2%) were diagnosed as underweight, and obesity was diagnosed in 91 males (10.9%) and 18 females (4.2%).

**Table 1.** Characteristics of study subjects.

|  | Male<br>(n=838) | Female<br>(n=426) |
| --- | --- | --- |
| Age (year) | 18.5 ± 0.6 | 18.3 ± 0.5 |
| Height (m) | 1.71 ± 0.06 | 1.58 ± 0.05 |
| Body weight (kg) | 62.2 ± 9.8 | 51.4 ± 6.8 |
| BMI (kg/m<sup>2</sup>) | 21.2 ± 3.0 | 20.5 ± 2.3 |
| Total fat mass (kg) | 11.2 ± 5.3 | 14.9 ± 4.5 |
| Body fat percentage (%) | 17.3 ± 5.6 | 28.4 ± 4.9 |
| Total muscle mass (kg) | 48.4 ± 5.3 | 34.5 ± 3.0 |
| ASM (kg) | 23.3 ± 2.9 | 16.1 ± 1.7 |
| SMI (kg/m<sup>2</sup>) | 7.9 ± 0.7 | 6.4 ± 0.5 |
| Total bone mass (kg) | 2.7 ± 0.3 | 2.0 ± 0.3 |
| Body water content (kg) | 34.5 ± 4.2 | 25.6 ± 2.5 |
| HGS (kg) | 38.6 ± 6.0 | 25.8 ± 4.3 |
| Energy intake (kcal/day) | 2,045 ± 695 | 1,562 ± 489 |
| Total protein intake (g/day) | 76.8 ± 31.2 | 60.3 ± 22.6 |
| Animal protein intake (g/day) | 44.7 ± 24.4 | 35.4 ± 16.9 |
| Vegetable protein intake (g/day) | 32.1 ± 11.7 | 24.9 ± 8.6 |
| Total fat intake (g/day) | 61.6 ± 23.3 | 50.5 ± 18.3 |
| Animal fat intake (g/day) | 29.7 ± 14.5 | 23.5 ± 10.5 |
| Vegetable fat intake (g/day) | 32.0 ± 11.5 | 27.0 ± 10.0 |
| Carbohydrate intake (g/day) | 286.6 ± 109.1 | 210.8 ± 71.9 |
| Frequency of breakfast intake | 7.2 ± 2.5 | 7.4 ± 2.4 |
| Physical activity (MET-minutes/week) | 2,091 ± 2351 | 1,371 ± 1638 |

BMI, body mass index; ASM, appendicular skeletal muscle mass; SMI, skeletal muscle mass index; HGS, handgrip strength; MET, metabolic equivalent. Data are expressed as mean ± standard deviation.

**Fig 2.**
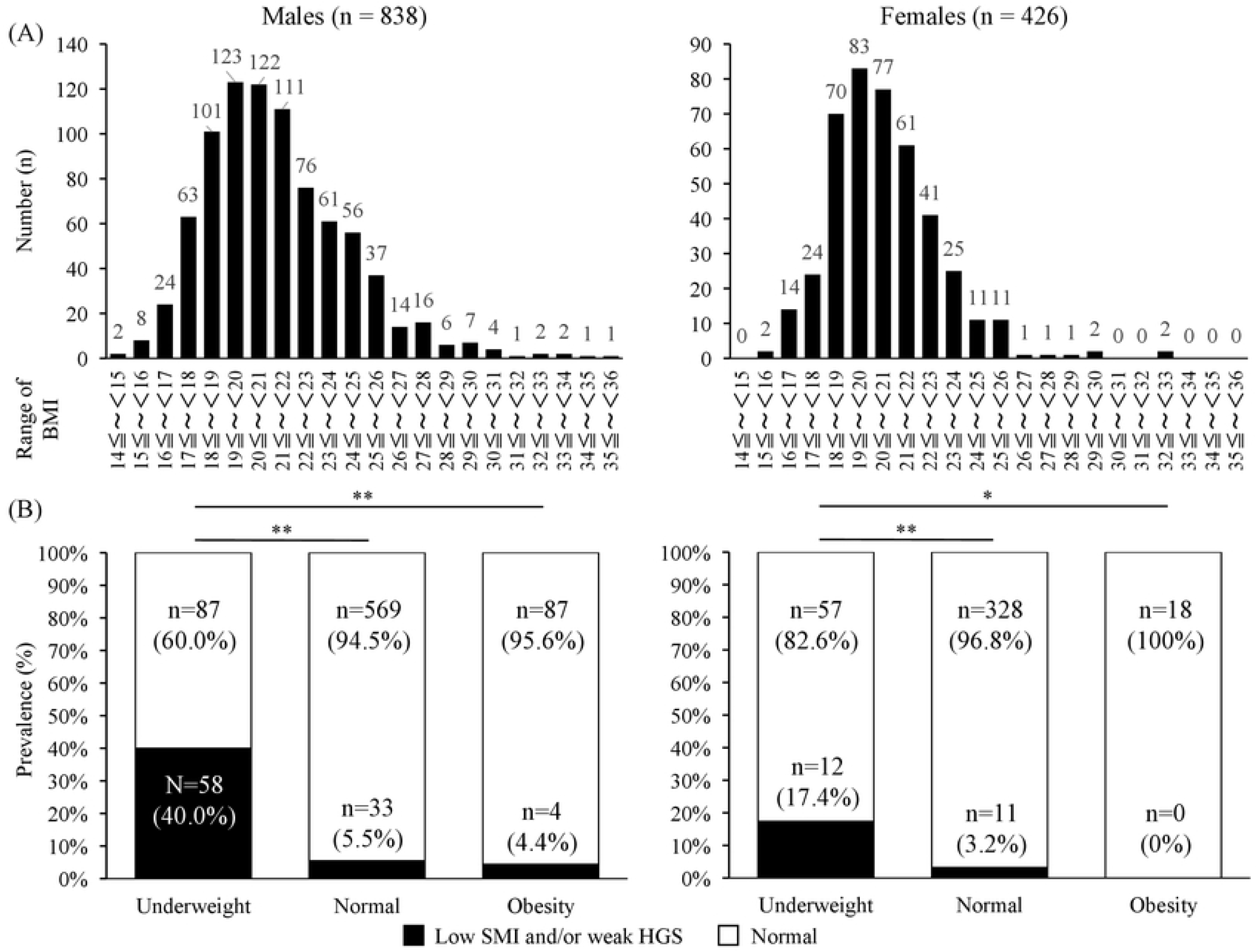
Body physique of university students. (A)Distribution of BMI and (B) prevalence of low SMI and/or weak HGS in each body physique in male and female university students. The prevalence was compared among the three groups (underweight, normal, and obesity) using the χ2-test. Then, as a post hoc test, multiple comparisons were performed using Tukey’s WSD test. **, *P* < 0.01.

The average SMIs in the males and females were 7.9 ± 0.7 and 6.4 ± 0.5 kg/m^2^, respectively, and both values were above the low SMI cutoff values. Low SMI was diagnosed in 75 males (8.9%) and 18 females (4.2%). The average HGS values in the males and females were 38.6 ± 6.0 and 25.8 ± 4.3 kg, respectively, and both values were above the weak HGS cutoff values. Weak HGS was diagnosed in 28 males (3.3%) and 10 females (2.3%). Then, sarcopenia was diagnosed in 8 males (1.0%) and 5 females (1.2%). There was a significantly higher prevalence of low SMI and/or weak HGS in the underweight individuals (40.0% in males and 17.4% in females) compared with individuals in the other BMI ranges in both sexes (Fig 2B).

### Simple linear regression analyses of the factors associated with SMI and HGS

The simple linear regression analyses indicated that SMI and HGS were positively associated with one another in both sexes (males, r = 0.53, *P* < 0.001; females, r = 0.53, *P* < 0.001) (Table 2). Furthermore, height, body weight, BMI, total fat mass, body fat percentage, total muscle mass, ASM, total bone mass, body water content, and physical activity were significantly associated with SMI and HGS in males (Table 2). Conversely, height, body weight, BMI, total fat mass, total muscle mass, ASM, total bone mass, and body water content were significantly associated with SMI and HGS in females (Table 2). In particular, SMI was positively correlated with energy intake and all nutrients, total/animal/vegetable protein, total/animal/vegetable fat, and carbohydrates, in females (Table 2).

**Table 2.** Simple linear regression analyses of the factors associated with SMI and HGS.

|  | SMI |  |  |  |
| --- | --- | --- | --- | --- |
|  | Male |  | Female |  |
|  | *r* | *P* value | *r* | *P* value |
| Age (year) | 0.01 | 0.774 | 0.00 | 0.930 |
| Height (m) | 0.19 | < 0.001 | 0.15 | 0.003 |
| Body weight (kg) | 0.70 | < 0.001 | 0.56 | < 0.001 |
| BMI (kg/m<sup>2</sup>) | 0.68 | < 0.001 | 0.56 | < 0.001 |
| Total fat mass (kg) | 0.37 | < 0.001 | 0.30 | < 0.001 |
| Body fat percentage (%) | 0.19 | < 0.001 | 0.07 | 0.143 |
| Total muscle mass (kg) | 0.88 | < 0.001 | 0.76 | < 0.001 |
| ASM (kg) | 0.86 | < 0.001 | 0.77 | < 0.001 |
| SMI (kg) | - | - | - | - |
| Total bone mass (kg) | 0.87 | < 0.001 | 0.76 | < 0.001 |
| Body water content (kg) | 0.95 | < 0.001 | 0.86 | < 0.001 |
| HGS (kg) | 0.53 | < 0.001 | 0.53 | < 0.001 |
| Energy intake (kcal/day) | 0.06 | 0.064 | 0.15 | 0.002 |
| Total protein intake (g/day) | 0.06 | 0.111 | 0.14 | 0.003 |
| Animal protein intake (g/day) | 0.04 | 0.228 | 0.12 | 0.017 |
| Vegetable protein intake (g/day) | 0.06 | 0.082 | 0.15 | 0.002 |
| Total fat intake (g/day) | 0.06 | 0.102 | 0.15 | 0.002 |
| Animal fat intake (g/day) | 0.07 | 0.037 | 0.14 | 0.003 |
| Vegetable fat intake (g/day) | 0.02 | 0.489 | 0.12 | 0.011 |
| Carbohydrate intake (g/day) | 0.06 | 0.103 | 0.12 | 0.013 |
| Frequency of breakfast (9 levels) | 0.05 | 0.153 | 0.09 | 0.068 |
| Physical activity (MET-minutes/week) | 0.17 | < 0.001 | 0.07 | 0.174 |

**Table 2. (continued)**
|  | HGS |  |  |  |
| --- | --- | --- | --- | --- |
|  | Male |  | Female |  |
|  | *r* | *P* value | *r* | *P* value |
| Age (year) | 0.04 | 0.231 | -0.04 | 0.412 |
| Height (m) | 0.36 | < 0.001 | 0.33 | < 0.001 |
| Body weight (kg) | 0.42 | < 0.001 | 0.41 | < 0.001 |
| BMI (kg/m<sup>2</sup>) | 0.29 | < 0.001 | 0.28 | < 0.001 |
| Total fat mass (kg) | 0.19 | < 0.001 | 0.21 | < 0.001 |
| Body fat percentage (%) | 0.07 | 0.030 | 0.06 | 0.189 |
| Total muscle mass (kg) | 0.56 | < 0.001 | 0.57 | < 0.001 |
| ASM (kg) | 0.59 | < 0.001 | 0.57 | < 0.001 |
| SMI (kg) | 0.53 | < 0.001 | 0.53 | < 0.001 |
| Total bone mass (kg) | 0.56 | < 0.001 | 0.58 | < 0.001 |
| Body water content (kg) | 0.55 | < 0.001 | 0.56 | < 0.001 |
| HGS (kg) | - | - | - | - |
| Energy intake (kcal/day) | 0.01 | 0.762 | 0.01 | 0.790 |
| Total protein intake (g/day) | 0.01 | 0.703 | 0.03 | 0.548 |
| Animal protein intake (g/day) | 0.02 | 0.614 | 0.02 | 0.631 |
| Vegetable protein intake (g/day) | 0.00 | 0.972 | 0.03 | 0.529 |
| Total fat intake (g/day) | 0.01 | 0.743 | 0.03 | 0.474 |
| Animal fat intake (g/day) | 0.03 | 0.343 | 0.03 | 0.480 |
| Vegetable fat intake (g/day) | -0.02 | 0.597 | 0.03 | 0.568 |
| Carbohydrate intake (g/day) | 0.01 | 0.823 | 0.00 | 0.932 |
| Frequency of breakfast (9 levels) | 0.03 | 0.340 | 0.05 | 0.291 |
| Physical activity (MET-minutes/week) | 0.13 | < 0.001 | 0.00 | 0.936 |

BMI, body mass index; ASM, appendicular skeletal muscle mass; SMI, skeletal muscle mass index; HGS, handgrip strength; MET, metabolic equivalent.

### Multiple linear regression analyses of the factors associated with SMI and HGS

To further examine the factors associated with SMI and HGS, we performed multiple linear regression analyses that included BMI, energy intake, frequency of breakfast intake, and physical activity as independent factors (Table 3). These variables were chosen because BMI, energy intake, and physical activity were significantly correlated with SMI and HGS in both sexes (Table 2). Additionally, frequency of breakfast intake has been reported to be associated with various health problems such as weight gain, glycemic control in diabetic patients, and the onset of proteinuria [22-24], and it also tended to be associated with SMI in females in the present study (*r* = 0.09, *P* = 0.068) (Table 2).

**Table 3.** Multiple linear regression analyses of the factors associated with SMI and HGS.

| SMI |  | BMI<br>(kg/m<sup>2</sup>) | Energy intake<br>(kcal/day) | Frequency of<br>breakfast<br>intake | Physical activity<br>(MET-minutes/week) |
| --- | --- | --- | --- | --- | --- |
| Model 1 |  |  |  |  |  |
| Male | $\beta$ | 0.68 | 0.03 | - | - |
| | $P$ value | < 0.001 | 0.232 | - | - |
| Female | $\beta$ | 0.56 | 0.14 | - | - |
| | $P$ value | < 0.001 | < 0.001 | - | - |
| Model 2 |  |  |  |  |  |
| Male | $\beta$ | 0.68 | - | 0.02 | - |
| | $P$ value | < 0.001 | - | 0.47 | - |
| Female | $\beta$ | 0.56 | - | 0.11 | - |
| | $P$ value | < 0.001 | - | 0.009 | - |
| Model 3 |  |  |  |  |  |
| Male | $\beta$ | 0.67 | - | - | 0.13 |
| | $P$ value | < 0.001 | - | - | < 0.001 |
| Female | $\beta$ | 0.56 | - | - | 0.06 |
| | $P$ value | < 0.001 | - | - | 0.138 |
| Model 4 |  |  |  |  |  |
| Male | $\beta$ | 0.67 | 0.01 | 0.02 | 0.13 |
| | $P$ value | < 0.001 | 0.736 | 0.556 | < 0.001 |
| Female | $\beta$ | 0.56 | 0.12 | 0.09 | 0.05 |
| | $P$ value | < 0.001 | 0.002 | 0.032 | 0.194 |

**Table 3. (continued)**
| HGS |  | BMI<br>(kg/m<sup>2</sup>) | Energy intake<br>(kcal/day) | Frequency<br>of breakfast<br>intake | Physical activity<br>(MET-minutes/week) |
| --- | --- | --- | --- | --- | --- |
| Model 1 |  |  |  |  |  |
| Male | $\beta$ | 0.29 | 0 | - | - |
|  | *P* value | < 0.001 | 0.904 | - | - |
| Female | $\beta$ | 0.28 | 0.01 | - | - |
|  | *P* value | < 0.001 | 0.869 | - | - |
| Model 2 |  |  |  |  |  |
| Male | $\beta$ | 0.29 | - | 0.02 | - |
|  | *P* value | < 0.001 | - | 0.55 | - |
| Female | $\beta$ | 0.28 | - | 0.06 | - |
|  | *P* value | < 0.001 | - | 0.201 | - |
| Model 3 |  |  |  |  |  |
| Male | $\beta$ | 0.29 | - | - | 0.11 |
|  | *P* value | < 0.001 | - | - | < 0.001 |
| Female | $\beta$ | 0.28 | - | - | 0 |
|  | *P* value | < 0.001 | - | - | 0.988 |
| Model 4 |  |  |  |  |  |
| Male | $\beta$ | 0.28 | -0.03 | 0.02 | 0.12 |
|  | *P* value | < 0.001 | 0.398 | 0.491 | < 0.001 |
| Female | $\beta$ | 0.28 | 0 | 0.06 | 0 |
|  | *P* value | < 0.001 | 0.982 | 0.206 | 0.989 |

BMI, body mass index; ASM, appendicular skeletal muscle mass; SMI, skeletal muscle mass index; HGS, handgrip strength; MET, metabolic equivalent; $\beta$ : standardized regression coefficient. Model 1 was adjusted for BMI and energy intake, Model 2 was adjusted for BMI and frequency of breakfast, Model 3 was adjusted for BMI and physical activity and Model 4 was adjusted for BMI, energy intake, frequency of breakfast and physical activity.

First, we constructed models 1, 2, and 3, which were adjusted for BMI because BMI is a major regulator of skeletal muscle mass and strength (Table 3). In males, we found significant positive correlations of physical activity with SMI and HGS after adjusting for BMI (*β* = 0.13, *P* < 0.001; *β* = 0.11, *P* < 0.001, respectively) (Table 3). In females, we found significant positive correlations of energy intake and frequency of breakfast intake with SMI after adjusting for BMI (*β* = 0.14, *P* < 0.001; *β* = 0.11, *P* = 0.009, respectively) (Table 3). In females, HGS was positively correlated only with BMI (Table 3).

Next, we constructed a model that included all factors (model 4). In males, we found that BMI and physical activity independently exhibited significant positive correlations with both SMI and HGS (SMI, *β* = 0.67, *P* < 0.001, *β* = 0.13, *P* < 0.001; HGS, *β* = 0.28, *P* < 0.001, *β* = 0.12, *P* < 0.001) (Table 3). In females, we found that BMI, energy intake, and frequency of breakfast intake independently exhibited significant positive correlations with SMI (*β* = 0.56, *P* < 0.001; *β* = 0.12, *P* = 0.002; *β* = 0.09, *P* = 0.032, respectively) and that only BMI independently exhibited a significant positive correlation with HGS (*β* = 0.28, *P* < 0.001) (Table 3).

## Discussion

The present study revealed that there was a higher proportion of underweight individuals than that of obese individuals; specifically, there were 1.6 times more underweight males and 3.8 times more underweight females (Fig 2A). It is feared that such underweight individuals may suffer from health problems in the future, such as sarcopenia, osteoporosis, and type 2 diabetes, which may shorten healthy life expectancy. Furthermore, this study revealed that in addition to BMI, energy intake, frequency of breakfast intake, and physical activities were significantly associated with skeletal muscle mass and strength in youth. Therefore, sarcopenia in youth was considered a part of secondary sarcopenia, that is, nutrition- and activity-related sarcopenia.

The increasing prevalence of underweight among young females has been a growing problem in Japan [25, 26]. However, this study showed that there is a high prevalence of underweight in males besides females. Additionally, high prevalence of low SMI and/or weak HGS were observed in underweight individuals of both sexes (Fig 2B). To prevent the onset of sarcopenia in later life, it is important to increase muscle mass and strength during youth and young adulthood (up to ∼40 years of age) and to maintain these factors beyond the age of 50 years because muscle mass and strength decrease with age [8]. Therefore, the results of this study suggest that the prevalence of sarcopenia may increase in the future and that we must be alert to this possibility.

The present study showed that the prevalence of weak HGS was low compared with that of low SMI (3.3% vs. 8.9% in males and 2.3% vs. 4.2% in females, respectively), resulting in a low prevalence of sarcopenia (1.0% in males and 1.2% in females) in both sexes. Goodpaster et al. reported that the decline in leg strength is much more rapid than the concomitant loss of muscle mass with aging [27]. This discrepancy may be due to the difference in muscle quality (MQ). MQ is defined as muscle strength per unit of muscle mass [8]. MQ considers to be affected by intramuscular fat infiltration and muscle fibrosis [28]. Taken together, HGS might have been preserved because the MQ was good in the young subjects with low SMI. In the future, histological investigation of skeletal muscle in the young subjects with low SMI, weak HGS, and sarcopenia is needed.

The present study showed that HGS was positively correlated with SMI in both sexes (Table 2). Furthermore, the factors significantly associated with HGS and those with SMI were almost the same (Table 2). These results suggests that muscle mass is a strong determinant of muscle strength. However, the correlation between HGS and SMI is not as strong (males, r = 0.53; females, r = 0.53). This is also because MQ is considered to vary not only in the elderly but also in the young. To date, there is no standard method for assessing MQ [8]. We recently reported that phase angle from bioelectrical impedance analysis can be a useful indicator of MQ [29]. The PhA declines with aging and subjects with sarcopenia have a lower PhA compared with those without it [29]. The relationship between the PhA and histological changes in skeletal muscle should be clarified in the future.

Interestingly, in addition to BMI, physical activity in males and energy intake in females were independently associated with SMI. The question then arises as to why there were sex differences in the factors independently associated with SMI. Muscle protein synthesis (MPS) is reported to be correlated with energy intake, particularly the branched-chain amino acid leucine [14]. According to the 2019 National Health and Nutrition Survey in Japan, the average energy intake of the females in the present study was much lower than that of elderly females over 75 years old (1,562 vs. 1,674 kcal) [1]. Conversely, the average energy intake of the males in the present study was higher than that of elderly males over 75 years old (2,045 vs. 2,008 kcal) [1], which suggests that the energy intake was considered sufficient for MPS in young males. Taken together, energy intake may be independently associated with SMI if it is insufficient for MPS. Conversely, physical activity may be independently associated with SMI if energy intake is sufficient for MPS. Inadequate protein intake and lack of exercise habits are thought to predispose people to sarcopenia and frailty [30, 31]. Furthermore, the effects of protein intake and physical activity on MPS have been reported to be stronger in the young than in the elderly [14, 15]. Therefore, adequate protein intake and exercise from a youthful age are necessary to prevent the development of sarcopenia in the future.

In the present study, frequency of breakfast intake was independently associated with SMI and HGS in females. According to the 2019 National Health and Nutrition Survey in Japan, both males and females in their twenties had the highest rate of skipping breakfast (15.0% in males and 10.2% in females) [1]. Increase in breakfast intake was associated with greater overall intake in subjects [32]. Additionally, the consumption of a moderate amount of protein at each meal stimulated 24-h MPS more effectively than skewed protein intake [33]. It has also been reported that skipping breakfast can induce stress and risk the appearance of proteinuria [24]. Taken together, regular breakfast intake is considered important for preventing sarcopenia.

This study has a few limitations. First, our study was cross-sectional in design and conducted in two universities of the Keiji area; thus, it could only examine associations among SMI, HGS, energy intake, frequency of breakfast intake, and physical activity. Second, dietary habits and physical activity were evaluated using questionnaires. Precise evaluation using objective indicators is necessary. First-year university students may have had special dietary habits and physical activity influenced by studying for entrance exams. To resolve these limitations, prospective studies are necessary in the future.

In conclusion, first-year university students showed a high incidence of being underweight with low SMI and/or weak HGS, but the prevalence of sarcopenia was low in both sexes. These young subjects may develop not only sarcopenia but also osteoporosis and type 2 diabetes in the future. BMI, energy intake, frequency of breakfast intake, and physical activity were factors independently associated with skeletal muscle mass and strength. It is necessary to provide early warnings and diet and exercise interventions for youth to prevent the development of sarcopenia.

## Data Availability

All relevant data are within the manuscript and its Supporting Information files.

## Acknowledgments

The authors would like to thank Hiromi Kusakabe for her secretarial assistance at the Clinical Research Institute, National Hospital Organization Kyoto Medical Center, Risa Hirai, and Yumiko Yamaguchi for their measurement assistance at the Kyoto Institute of Technology, Chikako Sugimoto, Yuko Imamura, Yumie Sakamoto, Hiromi Kato, and Junko Yamamoto for their measurement assistance at the Shiga University, and Enago (www.enago.jp) for the English language review.

## Author Contributions

**Conceptualization:** Toru Kusakabe, Hiroshi Arai, Yuji Yamamoto, Kazuwa Nakao

**Data curation:** Toru Kusakabe, Hiroshi Arai, Yuji Yamamoto, Yasunori Akamatsu

**Formal analysis:** Toru Kusakabe, Yasunori Akamatsu

**Funding acquisition:** Toru Kusakabe

**Investigation:** Toru Kusakabe, Yasunori Akamatsu

**Methodology:** Toru Kusakabe, Yasunori Akamatsu

**Project administration:** Toru Kusakabe, Hiroshi Arai, Yuji Yamamoto

**Resources:** Toru Kusakabe

**Supervision:** Toru Kusakabe, Kazuwa Nakao,

**Writing – original draft Preparation:** Toru Kusakabe

**Writing – Review & Editing:** Hiroshi Arai, Yuji Yamamoto, Kazuwa Nakao, Yasunori Akamatsu, Yuki Ishihara, Tetsuya Tagami, Akihiro Yasoda, Noriko Satoh-Asahara

## Funding

This study was supported in part by Grant-in-Aid for Clinical Research from the National Hospital Organization to T.K. (H30-NHO-03, R3-NHO-01) and Grant-in-Aid for Scientific Research (C) to T.K. (JSPS KAKENHI Grant No. 21K11691). This study was also supported in part by a grant from Issikai Association to T.K. The funders played no role in the study design, data collection and analysis, decision to publish, or preparation of the manuscript.

## Conflict of interest

The authors declare they have no conflict of interest with respect to this research study and paper.

## Notes

### Competing Interest Statement

The authors have declared no competing interest.

### Author Declarations

Consent to participate was obtained from all participants before study inclusion. This study was reviewed and approved by the Ethics Committee of National Hospital Organization Kyoto Medical Center (approval number: 18-106) and was conducted in compliance with the ethical principles stated in the Declaration of Helsinki.

